# Knowledge, Attitude, and Practices of Community Pharmacists on Antibiotic Resistance and Antimicrobial Stewardship in Lusaka, Zambia

**DOI:** 10.1101/2020.08.27.20181826

**Authors:** Steward Mudenda, Mutinta Hankombo, Zikria Saleem, Mohammad Jaffar Sadiq, Michelo Banda, Derick Munkombwe, Chiluba Mwila, Maisa Kasanga, Annie Chibwe Zulu, Jimmy Mapenzi Hangoma, Webrod Mufwambi, Lungwani Tyson Muungo, Ronald Mutati Kampamba, Andrew Munkuli Bambala, Noor Mohammed Abdulrahman, Muhammad Akram, John Bwalya Muma

## Abstract

Antibiotic resistance is among the major threats to global health. Due to limited information on the subject matter in Zambia, we assessed the knowledge, attitude, and practices of community pharmacists on antibiotic resistance and antimicrobial stewardship. We conducted a descriptive cross-sectional study among 144 randomly selected community pharmacists through a structured questionnaire. Data were analysed using Statistical Package for Social Sciences version 22 at 95% confidence level. A response rate of 91% (n=144) was achieved. The majority (63%) of the community pharmacists were male and were aged between 30 and 39 years. Interestingly, 93.8% had good knowledge while 67% had positive attitudes. Conversely, 75% of the community pharmacists demonstrated poor practices towards antibiotic resistance and antimicrobial stewardship. Even though community pharmacists had good knowledge and positive attitudes, they demonstrated poor practices that require quick educational interventions. There is a need to promote antimicrobial stewardship sensitization programmes among community pharmacists.

## Background

The decline in the introduction of new antibiotics on the market in the past decades has posed a great threat to the treatment of infectious diseases [1]. Antibiotic resistance is currently a global health issue that calls for urgent attention because effective treatment of infectious diseases is an essential component of both human and animal health management [2-4]. The impact of antibiotic resistance includes increased resource utilization, clinical or economic burdens, and increased use of broad-spectrum antibiotics, increased morbidity, and mortality [5]. Although antibiotic resistance is an inevitable consequence of antibiotic use, the rate and extent of propagation of resistant bacteria are governed by human behavior such as excessive and inappropriate use of antibiotics in health systems [6]. Inappropriate use of antibiotics is directly related to the tendency towards self-medication and the unnecessary use of antibiotics for viral diseases [7].

It has been observed that most people suffering from viral upper respiratory tract infections such as common colds tend to misuse antibiotics for treatment [8, 9]. This irrational use of antibiotics for some respiratory tract diseases escalates cases of antibiotic-resistant bacteria [10]. Being among respiratory tract infections, Coronavirus Disease 2019 (COVID-19) is likely to escalate antibiotic resistance due to an expected over-prescribing, inappropriate use, and self-prescription of antibiotics [11-14]. Strategies must be implemented for the management of upper respiratory tract infections and reduce the inappropriate consumption of antibiotics [15]. Therefore, prescribers and all healthcare workers need to be watchful as they prescribe and dispense antibiotics even in times of global pandemics.

Community pharmacists (CPs) are the first point of contact for community members seeking pharmaceutical and medical services due to their ease of accessibility, less waiting time, and cheaper costs [16-19]. Nevertheless, members of the community find it easier to access the CPs and acquire their medical help [20]. It is noteworthy that members of the community trust the CPs and thus obtain their antibiotics through interaction with CPs [21]. In developing countries, there has been an increase in the use of non-prescribed antibiotics which has been exacerbated by non-prescription sales of antibiotics in community pharmacies [22, 23]. Global estimations have indicated that more than 50% of antibiotics are accessed privately without a prescription; with increased evidence indicating that CPs are failing to promote the rational and appropriate use of antibiotics [24-26].

The supply of antibiotics from a community pharmacy without a prescription usually involves interaction with a community pharmacist [27, 28]. The problem of dispensing antibiotics without a prescription has been reported in low-and middle-income countries [29, 30]. Therefore, changing the publics’ attitudes and improving their knowledge regarding antibiotic use is the responsibility of the community pharmacist who happens to be the first point of contact and source of these antibiotics [31]. Emphasis has therefore been placed on improving the role of the pharmacist as the main supplier and regulator of antibiotics in communities [32].

Being a global health problem, it is therefore important that CPs have good knowledge, positive attitude, and good practices (KAP) towards antibiotic resistance [33, 34]. However, solving the problem of antibiotic resistance requires that health systems initiate and implement Antimicrobial Stewardship Programmes (ASPs) [35, 36]. CPs may play a crucial role in the fight against antimicrobial resistance by acting as proponents of ASPs [37-39]. ASPs promote the rational use of antibiotics and reduce antibiotic resistance [40, 41]. As a member of the medical community, CPs must act positively in the fight against antibiotic resistance by embracing and participating in ASPs [42-44].

In the Zambian setup, there is limited information on the knowledge, attitude, and practices of community pharmacists on antibiotic use and resistance, and ASPs. Therefore, this study is very beneficial to the Zambian pharmacists and all pharmacists globally. This study is also of benefit to other healthcare workers because antibiotic resistance affects everyone. CPs should promote the rational use of antibiotics even in times of global health pandemics to prevent the worsening of the emergence of antibiotic-resistant bacteria.

## Methods

### Study Design and Site

This was a descriptive cross-sectional study that was conducted in community pharmacies in Lusaka Zambia from March 2019 to August 2019. Lusaka is the Capital city of Zambia and has more than 50% of registered community pharmacies in Zambia [45]. The study involved registered CPs who provided consent to participate in the study. Pharmacy technologists, dispensers, unregistered pharmacies, and drug stores were excluded from this study.

### Sample size

The sample size was calculated using the Slovin’s formula [46]. The sample size calculation was done at a 95% confidence level and a 5% margin of error. However, a 10% surplus was added to the sample size to overcome challenges associated with non-responses and incomplete responses. The community pharmacists were chosen using simple random sampling from a total of 225 registered community pharmacies in Lusaka at the time of the study as was obtained from the Zambia Medicines Regulatory Authority (ZAMRA) Register [45].

### Data collection tool: Structure self-administered questionnaire

Data collection was done using a structured self-administered questionnaire. The questionnaire was adopted and modified from similar studies [33, 47]. The questionnaire was pre-validated for its simplicity, accuracy, clarity, understandability, and relevance by experts in the field of pharmacy. This gave room for modification of the initial questionnaire. Later, the questionnaire was pretested among 12 community pharmacists for its consistency. Then the final structured questionnaire was used to collect data on sociodemographic characteristics, knowledge, attitude, and practices of community pharmacists on antibiotic resistance and AMS. The responses on knowledge and attitude questions were measured using a 5-point Likert-scale as follows; 1= ‘strongly disagreed’, 2= ‘disagreed’, 3= ‘neutral’, 4= ‘agreed’, and 5= ‘strongly agreed’. The responses on practice questions were measured using a Likert scale as follows; 1= ‘never’, 2= ‘rarely’, 3= ‘occasionally’, 4= ‘often’, and 5= ‘always’. Reverse coding was done for negative worded questions. Scores for knowledge ranged from 5 to 25, scores equal to or above 12 indicated good knowledge while scores below 12 indicated poor knowledge. Scores for attitude ranged from 6 to 30, scores equal to or above 15 indicated positive attitude while scores below 15 indicated negative attitude. Scores for practices ranged from 7 to 35, scores equal to or above 17 indicated good practices while scores below 17 indicated poor practices. Dependent variables included knowledge, attitude, and practices. Independent variables included age, sex, level of education, and work experience.

### Data analysis

The data were analysed using Statistical Package for the Social Sciences (SPSS) version 22.0. Data was initially entered into the Microsoft Excel spread sheet before being exported to SPSS. Data was presented in form of tables and charts and a Likert scale was used to determine the level of knowledge, attitude, and practices. A p<0.05 was used to indicate statistical significance at a 95% confidence level. Fisher’s exact test was used to determine the relationship between categorical variables.

### Ethical approval

The study was approved by the University of Zambia Health Sciences Research Ethics Committee (UNZAHSREC), protocol ID: 20190217024, IORG no: 0009227, IRB no: 00011000. All participants provided consent by reading and signing the consent form prior to responding to the questionnaire.

## Results

### Demographic characteristics of participants

The demographic characteristics of 144 community pharmacists (CPs) who took part in the study are shown in Table 1. Most of the respondents were male (63%) and were aged between 30 and 45 years (65%).

**Table 1:** Characteristics of participants (n= 144)

| Variable | Category | Frequency (%) | p-value |
| --- | --- | --- | --- |
| Sex | Male | 91 (63) | 0.002 |
|  | Female | 53 (37) |  |
| Age | 20 - 29 | 28 (19) | <0.0001 |
|  | 30 - 39 | 94 (65) |  |
|  | 40 - 49 | 18 (13) |  |
|  | ≥ 50 | 4 (3) |  |
| Education level | Degree | 134 (93) | <0.0001 |
|  | Masters | 10 (7) |  |
| Work experience (years) | 1 – 4 | 54 (38) | <0.0001 |
|  | 5 – 9 | 64 (44) |  |
|  | ≥10 | 26 (18) |  |

### Overall knowledge, attitude, and practice scores on antibiotic resistance and AMS

The majority of the CPs 93.8% (n=135) had good knowledge and 67.4% had positive attitude with regards to antibiotic resistance. Unfortunately, 75.7% of the CPs had poor practice regarding antibiotic resistance and antimicrobial stewardship (Table 2).

**Table 2:** Overall knowledge, attitude, and practice scores on antibiotic resistance and AMS.

| Variable | Good (n, %) | Poor (n, %) |
| --- | --- | --- |
| Knowledge | 135 (93.8%) | 9 (6.2%) |
| Attitude | 97 (67.4%) | 47 (32.6%) |
| Practice | 35 (24.3%) | 109 (75.7%) |

### Community pharmacists’ knowledge of antibiotic resistance

The majority of the CPs (29.2% agreed, 67.4% strongly agreed) that antibiotic resistance occurs when antibiotic no longer work to treat bacterial infections. The majority of the CPs (29.7% agreed, 69.7% strongly agreed) that antibiotic resistance occurs due to misuse of antibiotics and it is public health problem in our communities (Table 3).

**Table 3:** Community pharmacists’ knowledge of antibiotic resistance.

| Variable | Responses |  |  |  |  |
| --- | --- | --- | --- | --- | --- |
|  | SD (%) | D (%) | N (%) | A (%) | SA (%) |
| Antibiotic resistance is when antibiotics no longer work to treat bacterial infections | 00 (0.0) | 3 (2.0) | 2 (1.4) | 42 (29.2) | 97 (67.4) |
| Misuse of antibiotics leads to a loss of sensitivity of an antibiotic to a specific pathogen | 00 (0.0) | 00 (0.0) | 00 (0.0) | 43 (29.9) | 101(70.1) |
| Antibiotics resistance is a problem in our community | 00 (0.0) | 00 (0.0) | 1 (0.7) | 36 (25) | 107(74.3) |
| Bacteria which are resistant to antibiotics can spread from person to person | 2 (1.4) | 5 (3.5) | 12 (8.3) | 38 (26.4) | 87 (60.4) |
| Many infections are becoming increasingly resistant to treatment using antibiotics | 00 (0.0) | 00 (0.0) | 8 (5.6) | 66 (45.8) | 70 (48.6) |
| SD=Strongly Disagreed; D=Disagreed; N=Neutral; A=Agree; SA=Strongly Agreed |  |  |  |  |  |

### Community pharmacists’ attitude towards antibiotic resistance

The majority of the CPs 69.4% agreed that antibiotic resistance is a public health problem. The majority of the CPs 57.6% agreed that appropriate use of antibiotics improves patient care and outcome (Table 4).

**Table 4:** Community pharmacists’ attitude towards antibiotic resistance.

| Variable | Responses |  |  |  |  |
| --- | --- | --- | --- | --- | --- |
|  | SD (%) | D (%) | N (%) | A (%) | SA (%) |
| Antibiotic resistance is a serious public health problem | 00 (0.0) | 00 (0.0) | 00 (0.0) | 100(69.4) | 44 (30.6) |
| Appropriate use of antibiotics can improve patient care and outcome | 00 (0.0) | 00 (0.0) | 00 (0.0) | 83 (57.6) | 61 (42.4) |
| Appropriate use of antibiotics can reduce problems of antibiotic resistance. | 00 (0.0) | 00 (0.0) | 00 (0.0) | 87 (60.4) | 57(39.6) |
| Adequate training on antibiotic resistance should be provided to retail pharmacists | 00 (0.0) | 8 (5.6) | 43(29.9) | 66 (45.8) | 27 (18.8) |
| Relevant conferences, workshops, and other educational activities are required to be attended by retail pharmacists to enhance understanding of antibiotic resistance. | 00 (0.0) | 16(11.1) | 38(26.4) | 64 (44.4) | 26 (18.1) |
| Pharmacists have a responsibility to take a prominent role in antibiotic resistance and infection-control programs in the health system. | 00 (0.0) | 00 (0.0) | 1 (0.7) | 56 (38.9) | 87 (60.4) |

**SD=Strongly Disagree; D=Disagree; N=Neutral; A=Agree; SA=Strongly Agree**

### Community pharmacists’ practice with regards to antimicrobial resistance and AMS

The majority of the CPs (32.6%) rarely collaborates with other healthcare workers in activities that promote infection control and AMS. In addition, the majority of the CPs do not take part in antibiotic awareness campaigns nor educate the public on antibiotic use (Table 5).

**Table 5:** Community pharmacists’ practice regarding antibiotic resistance and AMS.

| Variable | Responses |  |  |  |  |
| --- | --- | --- | --- | --- | --- |
|  | Never<br>(%) | Rarely<br>(%) | Occasionally<br>(%) | Often<br>(%) | Always<br>(%) |
| I dispense antibiotics on prescriptions with complete clinical information. | 00 (0.0) | 2 (1.4) | 00 (0.0) | 99 (68.7) | 43 (29.9) |
| I dispense antibiotics for durations longer than prescribed by the physician on patient request. | 47 (32.6) | 45 (31.3) | 41 (28.5) | 10 (6.9) | 1 (0.7) |
| I screen antibiotic prescriptions following local guidelines before dispensing. | 00 (0.0) | 3 (2.1) | 7 (4.9) | 51 (35.4) | 83 (57.6) |
| I collaborate with other health professionals on infection control and antimicrobial stewardship. | 7 (4.9) | 47 (32.6) | 43 (29.9) | 31 (21.5) | 16 (11.1) |
| I take part in antimicrobial-awareness campaigns to promote the optimal use of antimicrobials. | 8 (5.6) | 71 (49.3) | 36 (25.0) | 18 (12.5) | 11 (7.6) |
| I educate patients on the use of antibiotics and resistance-related issues. | 00 (0.0) | 56 (38.9) | 31 (21.5) | 23 (16.0) | 34 (23.6) |
| I dispense antibiotics without a prescription | 00 (0.0) | 6 (4.2) | 18 (12.5) | 112 (77.8) | 8 (5.5) |

### Relationships between sociodemographic characteristics and knowledge, attitude, and practices

Statistical significance was found between age and attitude, age and practices, work experience and knowledge, and work experience and practices (Table 6).

**Table 6:**
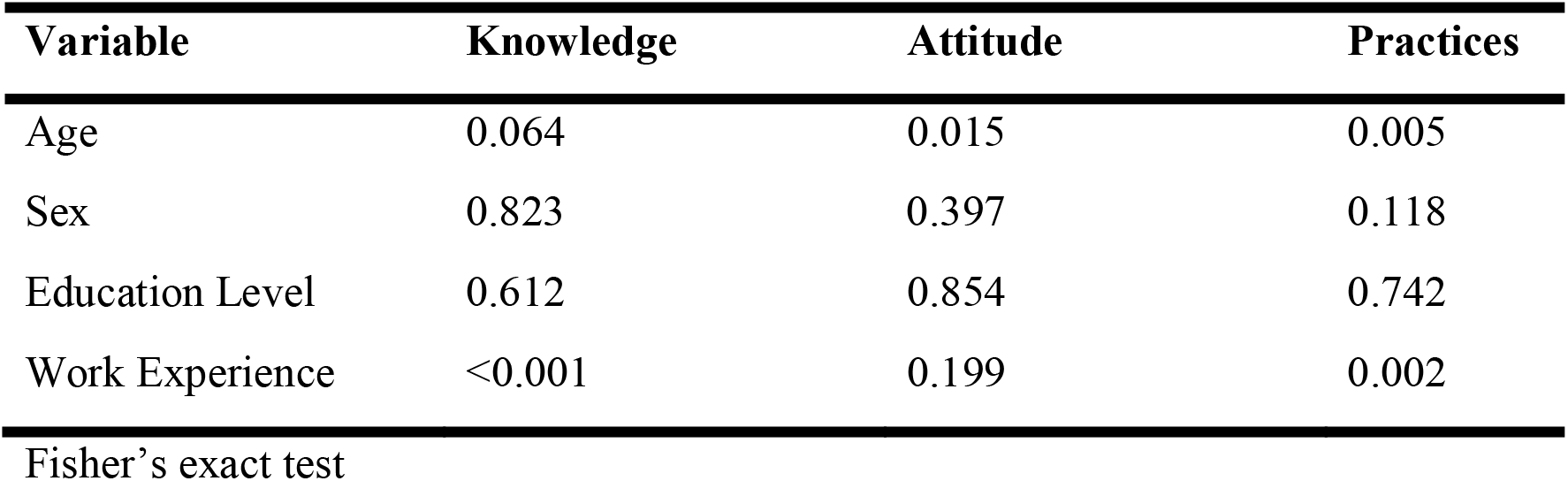
Relationship between sociodemographic characteristics and knowledge, attitude, and practice of the community pharmacists.

## DISCUSSION

We assessed that knowledge, attitude, and practice of community pharmacists (CPs) on antibiotic resistance and antimicrobial stewardship. Currently, there is limited information about the knowledge, attitude, and practice of CPs on antibiotic resistance and AMS in Zambia.

### Community pharmacists’ knowledge of antibiotic resistance

Our study found that 93.8% of CPs had good knowledge of antibiotic resistance that was related to their work experience. Similar results were reported in other studies [33, 48-53]. The similarities concerning knowledge about antibiotic resistance may be attributed to the adequate knowledge on antibiotics which pharmacists acquire during their university training and practice. Unfortunately, the implementation of the good knowledge may be very poor [53]. In Thailand, different results were reported which showed that most pharmacists had inadequate knowledge [54]. Good knowledge of antibiotic use and resistance prevents community pharmacists from irrational dispensing of antibiotics, and this is a progress in the fight against antibiotic resistance [55]. On the other hand, insufficient knowledge results in CPs dispensing antibiotics irrationally thereby exacerbating antibiotic resistance [56, 57].

In our study, the majority of the CPs strongly agreed that antibiotic resistance occurs when antibiotics fail to treat bacterial infections and that it usually occurs when there is overuse and misuse of antibiotics. Another study reported similar findings [47]. Many bacterial infections cannot be treated because of the misuse and overuse of antibiotics [58-63]. Antibiotic consumption has been linked to antibiotic resistance [64-67]. Therefore, CPs must help in the appropriate use of antibiotics to reduce on emergence of antibiotic-resistant bacteria. In addition, all healthcare systems must develop initiatives and solutions to reduce antibiotic consumption and antimicrobial resistance [68, 69].

The majority of the CPs in our study strongly agreed that antibiotic resistance is a public health problem in our communities. Similar findings were reported in Egypt [70]. Antibiotic resistance is a public health problem that will lead to increased morbidity and mortality if not controlled [64]. In the current study, the majority of the CPs strongly agreed that resistant bacteria can spread from one person to another and this makes infections difficult to treat. The World Health Organization (WHO) reported that resistant bacteria do spread from one person to another and infections caused by resistant bacteria are very difficult to treat [71]. We need to combat antibiotic-resistant bacterial infections and reduce morbidity and mortality associated with antimicrobial resistance [72].

### Attitudes of community pharmacists towards antibiotic resistance

Our study found that 67.4% of the CPs had a positive attitude towards antibiotic resistance that was related to their age. Our findings are encouraging because the positive attitude of CPs may influence them not to dispense antibiotics without a prescription [73]. In Syria, the majority of CPs had negative attitude towards antibiotic use and resistance [74]. The negative attitude influences CPs to dispense antibiotics without a prescription [59].

In the present study, the majority of the pharmacists agreed that antibiotic resistance is a serious public health problem. Similar results were reported in other studies [59, 75]. The majority of the community pharmacists in our study agreed that the appropriate use of antibiotics can reduce problems of antibiotic resistance. Appropriate use of antibiotics will help reduce the threat of antibiotic resistance [76]. It is therefore important that community pharmacists act as antibiotic stewards in the promotion of rational use and prescribing of antibiotics. Community pharmacists are supposed to promote the rational, economic prescribing and appropriate use of medicines as a means of good pharmacy practice [77]. Our findings show that the majority of the pharmacists agreed that adequate training, workshops, and other relevant activities on antibiotic resistance are recommended [78]. Relevant training for community pharmacists can help promote the rational use of antibiotics and thus help to curb antimicrobial resistance. Our study reported that community pharmacists working in collaboration with other healthcare providers must take prominent roles in infection control and antimicrobial stewardship programs. Therefore, the majority of our participants reported that they collaborated with other healthcare workers in infection control and antimicrobial stewardship programs. Other researchers also reported the same findings and concluded that collaboration among healthcare workers improves the rational use and prescribing of antibiotics [33, 79, 80].

### Practices community pharmacists towards antibiotic resistance and AMS

Our study revealed that the majority (75.7%) of CPs had a poor practice regarding antibiotic resistance and AMS. The younger and less experienced CPs showed poorer practices compared to the older and experienced CPs. In Pakistan, similar results were reported [33]. Poor practice towards antibiotic resistance calls for quick educational intervention programmes among CPs [81]. Their practice could be affected by lack of continuing education and other external factors.

Our study highlighted that the majority (68.8%) of the CPs often dispensed antibiotics on prescriptions with complete clinical information. Similar findings were reported in Albania and Tanzania respectively [82, 83]. CPs must ensure that they dispense antibiotics on prescriptions with full clinical information and counsel patients on how to use antibiotics [84]. Lack of providing full clinical information and counselling patients on antibiotic use may lead to the escalation of antibiotic resistance.

The majority of the CPs in our study never dispensed antibiotics for durations longer than prescribed by the prescriber or on patient request. Antibiotics should not be dispensed for prolonged durations because this practice can lead to abuse and misuse, which could result in antibiotic resistance [85]. The present study showed that most CPs always screen antibiotic prescriptions following the local guidelines before dispensing. CPs need to ensure that they validate and interpret the prescription correctly based on the recommended guidelines. This promotes the dispensing of appropriate medicines to the right patient [86, 87].

Our study revealed that the majority of the CPs rarely collaborated with other healthcare workers in infection control and ASPs. Hayat and colleagues reported similar findings [88]. This could be because most countries do not have well-defined ASPs for community pharmacy practice [33]. All countries are encouraged to start and steer successful ASPs [89]. In Ethiopia, Erku reported that the majority of the community pharmacists collaborated with other healthcare workers in infection control and ASPs [80]. Collaboration between pharmacists and physicians helps to reduce antibiotic resistance [90, 91]. The collaboration by pharmacists and physicians also helps pharmacists intervene in certain inappropriately prescribed antibiotics thus helping to reduce antibiotic resistance [92]. Also, pharmacists must unite in the fight against antibiotic resistance [93]. CPs must be encouraged to participate in ASPs because they play a vital role in regulating the use of antibiotics [94-96].

The majority of the CPs in our study rarely participated in antibiotic-awareness campaigns that promote the optimal use of antibiotics. Other studies found similar results [33, 97, 98]. As a result of this, the majority of the CPs rarely educated patients on the use of antibiotics and resistance-related issues. In contrast, a study in China reported that CPs usually educate patients on antibiotics and antibiotic resistance [99]. Patient education is very important in the provision of community pharmaceutical services and helps reduce infections caused by antibiotic-resistant bacteria [86, 100, 101].

All the CPs in our study revealed that they dispensed antibiotics without a prescription. Antibiotics, just like other prescription only drugs, must be dispensed on prescription only [102]. Dispensing antibiotics without prescription is very common in most countries in the world [83, 103-107]. This practice contributes greatly to the emergence of antibiotic-resistant bacteria [108-114]. In Zambia, it is a common practice of selling and dispensing antibiotics without a prescription [115]. This practice may escalate antibiotic resistance and thus requires urgent interventions. Therefore, pharmacists must continue playing the role of gatekeepers of antibiotics [116]. Educational interventions must be put in place to improve the appropriate prescribing and dispensing of antibiotics [117-119].

### Implications for future policy and practice

Concerning policy implications for this research study, antimicrobial stewardship programmes aimed at reducing antimicrobial resistance (AMR) need to be promoted in a multi-disciplinary approach. In Zambia, the National Action plan (NAP) on AMR was developed to address this gap through One Health approach [120]. Currently, the status of AMR in Zambia is mainly focussed on knowledge strengthening as well as evidence obtained from surveillance and research activities. Further training of community pharmacists in Zambia should involve bridging the gap between knowledge and practice. Substantial effort need to be directed towards motivating community pharmacists in undertaking on the job of antimicrobial training through continuous education programmes such as continuous pharmacy/medical education (CPE/CME) workshops [121]. This will be helpful as there is evidence that most pharmacists have not undertaken antimicrobial stewardship (AMS) trainings in Zambia [122]. The government of the republic of Zambia, other stakeholders, and cooperating partners need to institute inclusive policies for both private and public training institutions that will enable revision of training curriculum content on antimicrobial resistance which may subsequently contribute to reducing AMR. Further focus should be directed to private hospitals and clinics which are easily accessible to members of the public and might be the likely target in implementing AMS programmes. With this research submission, it is recommended that every state must inculcate a ‘rationalised antibiotic usage policy’ into their ‘national health policies’ to avoid any serious complications and outcomes in future. It should also be recommended the prescription and usage of antibiotics based on proofs but not empirical applications.

### Limitations of the study

We acknowledge that the current study has some limitations. It focused on assessing the knowledge, attitude, and practices of community pharmacist leaving out pharmacy technologists who work under the supervision of the pharmacists. Therefore, the results should be interpreted in the context of community pharmacists. Study was led in Lusaka province only; the results could not show the practice, knowledge, and attitude of community pharmacists working in further provinces of the Zambia. Since we did not include community pharmacists from all the ten provinces of Zambia, and so, generalization of the results should be done with caution. In addition, self-governed forms used in this research are disposed to common interest unfairness.

The study only focussed on registered community pharmacists and this could create a bias on the findings as illegal drug outlets are a nuisance in the country and could be the important drivers of antibiotic resistance and hamper the efforts towards AMS activities. In any case, this study has provided the much needed baseline information for future research work in as far as antimicrobial resistance in relation to community pharmacists is concerned.

### Conclusion

Community pharmacists in Lusaka Zambia had good knowledge and attitude but poor practices regarding antibiotic resistance. There is a need to promote ASPs in community pharmacies that may lead to the improved practice of community pharmacists with regards to antibiotic use, resistance, and AMS. Community pharmacists must be recognized as key players in the control and prevention of infections among community members. Therefore, educational intervention programmes with regards to antibiotic use, resistance, and AMS should be periodically focussed on community pharmacy practice.

## Data Availability

Data is available through links

## Acknowledgments

We would like to acknowledge the owners of the community pharmacies and the pharmacists for cooperating with us during data collection from their premises. We are grateful to the University of Zambia Library for providing access to many articles that were used for reference in this manuscript.

## Declaration of interest

### Notes on contributors

Steward Mudenda (MSc) is clinical pathologist and public health pharmacist, PhD student of infectious diseases, Lecturer and Researcher in the University of Zambia, Department of Pharmacy. Mutinta Hankombo (BPharm) is a hospital pharmacist at the University Teaching Hospitals in Lusaka, Zambia. Zikria Saleem (PhD) is an associate professor of Clinical Pharmacy in the University of Lahore, Pakistan. Mohammad Jaffar Sadiq (PhD) is an associate professor of Pharmacology in the Batterjee Medical College for Science and Technology in Jeddah, Saudi Arabia. Michelo Banda (MSc) is a Lecturer and Researcher of specialized of Basic Sciences in the University of Zambia, Department of Pharmacy. Derick Munkombwe (PhD) is a Lecturer and Researcher specialized in Pharmacology in the University of Zambia, Department of Pharmacy. Chiluba Mwila (PhD) is a Lecturer and Researcher of Pharmaceutics in the University of Zambia, Department of Pharmacy. Maisa Kasanga (MSc) is a Biomedical Scientist, PhD student of Public Health at Zhengzhou University, China. Annie Zulu (BPharm) is a hospital pharmacist at Levy Mwanawasa University Teaching Hospital, Zambia. Jimmy Mapenzi Hangoma (MSc) is a PhD student, Lecturer and Researcher of Clinical Pharmacy at Levy Mwanawasa Medical University. Webrod Mufwambi (MSc) is a PhD student, Lecturer and Researcher of Clinical Pharmacy in the University of Zambia, Department of Pharmacy. Lungwani Tyson Muungo (PhD) is a Lecturer and Researcher of Pharmaceutics at the University of Zambia, Department of Pharmacy. Ronald Mutati Kampamba (MSc) is a Lecturer and Researcher of Pharmacy Practice, Biopharmacy, and Medicinal Chemistry in the University of Zambia, Department of Pharmacy. Andrew Munkuli Bambala (MSc) is a clinical pharmacist specialized in Toxicology at the University Teaching Hospitals in Lusaka, Zambia. Noor Mohammed Abdulrahman (MSc) is a clinical pharmacist in the University of Basrah, College of Pharmacy, Iraq. Muhammad Akram (PhD) is an associate professor in the Faculty of Life Sciences, Government College University Faisalabad. John Bwalya Muma (PhD) is a professor of Public Health in the Department of Disease Control, University of Zambia.

